# Embracing Pressure. A qualitative analysis of team strengthening principles in military and elite sporting domains and consideration of their applicability to healthcare teams

**DOI:** 10.64898/2026.07.29.26356354

**Authors:** Peter McCulloch, Laurie Earl, Mark Sujan, Mudathir Ibrahim, Elizabeth Sutton

## Abstract

**Background:** The Crew Resource Management (CRM), teamwork training model widely used in healthcare improves teamwork behaviours, but evidence of improved patient outcomes is weak. Other professions with teams displaying high performance under pressure, such as military units and sporting teams may have lessons for teamworking in healthcare. We conducted a qualitative study with trainers from these environments, to explore aspects of their approaches and beliefs which might inform an alternative model for healthcare team strengthening.

**Methods:** We conducted and thematically analysed semi-structured interviews with 14 coaches/leaders of military and sporting teams. The findings were used to define principles for team strengthening programmes. Experienced healthcare staff and Human Factors experts reviewed these principles and identified challenges in translating them into healthcare environments.

**Results:** We identified 6 themes and 18 sub-themes. The principles identified encompassed (a) Deliberate self-development of team values by the group, their promotion by leaders and maintenance by experienced team members (b) Frequent intensive realistic whole team training, and full integration of sub-teams, (c) Celebration of success and encouragement of an attitude of embracing pressure (d) Use of Buddying and mentoring, symbols of belonging and team social events.

Important contextual issues requiring attention when adapting these principles for healthcare environments were identified.

**Conclusion:** The approaches to creating cultures of mutual supportiveness associated with high performance under pressure in military and sports teams were very similar. Adoption of these in healthcare may be feasible, but would require significant contextual adaptation. Pilot studies would allow refinement and evaluation of potential benefits.

## BACKGROUND

The potential for medical care to cause harm has become well recognised and studied over the last 30 years^1^. Harm directly caused by treatment has been quantified and analysed in considerable depth, but harms caused by delayed or suboptimal care are more difficult to evaluate, and may be equally important. Communication and collaboration failures are persistently identified as major contributors to safety incidents, so considerable attention has been paid to how team interactions could be improved. This attention to team function has formed part of the Human Factors approach to safety improvement which has become the dominant paradigm in this field. Human Factors touches on two distinct sets of issues which may contribute to patient harm. Systems of work may be inadequately adapted to human workload, cognitive and perceptual capacities, but the interaction between clinical team members may also contribute to risk via issues such as hierarchy, lack of psychological safety, or unhelpful inter-group relationships. In the latter sphere, the majority of efforts to improve team interaction have been based on the non-technical skills model developed by Helmreich^2^ and widely adopted by civil aviation. Unfortunately, the major reduction in avoidable harms envisaged by pioneers has not been realised over the last 20 years^3,4,5^, giving cause for re-evaluation of our approaches to intervention. The recognition that the options for improving team performance in healthcare may not have been comprehensively explored^6,7^ has led to an interest in learning from other high reliability industries, and from other settings in which consistent high performance under pressure is necessary for success.

Team training Programmes in healthcare based on the aviation Crew Resource Management (CRM) model^8,9^ have shown improvement in measures of team interaction, team member perceptions and team climate. Evidence of a positive effect on patient outcomes, however, is less robust. Systematic reviews consistently report improvement, but are biased by inclusion of the very large Veterans Administration MedTeams study^10^ whose findings are undermined by a design which compared units which had already received training and others which had not, in a programme where the criterion for priority in receiving training was “readiness”. We were therefore interested in revisiting different approaches to team strengthening, and included this amongst the interventions in our preparation for a large-scale study to develop and evaluate a multi-modal Human Factors intervention to improve rescue responses for critically ill patients following surgery. In a search for promising approaches to strengthening healthcare teams, we set out to identify experts with a proven ability to produce teams capable of communicating and cooperating at a high level under pressure. We identified elite sports coaches and professional military trainers as two groups with strong claims to expertise of this nature. We sought to explore their views on how teamwork can be developed and maintained in a qualitative interview-based study, aiming to define a set of core principles which could become the basis for an alternative theory of teamwork training in healthcare. The immediate purpose of the exercise was to develop proposals for a team strengthening intervention for our intervention study. Aware of the risks inherent in attempting to translate procedures and solutions between very different environments^1^, we asked a group of Healthcare Human Factors experts and a range of experienced frontline clinical staff members to review the applicability of our proposals in a healthcare environment, and modified them based on this analysis.

## METHODS

### INTERVIEWEE RECRUITMENT AND SAMPLING STRATEGY

We selected interviewees using a purposive sampling strategy which aimed to incorporate expertise from trainers of high-performing teams in military and sporting backgrounds, including some who were familiar with the healthcare environment. We used both direct and indirect approaches to recruit subjects, including an element of “snowballing” by asking for recommendations from interviewees. The main selection criterion was having coached a team or teams, either military or in team sports, with a record of excellent performance in a high-pressure, high-stakes environment.

A pilot interview with a prominent sports coach was used to refine a topic guide, clarify questions and interview strategy. A series of high-profile elite team sports coaches and leaders, and experts in military training were then approached. Participants gave written informed consent, and were asked for permission to use direct quotations, specifying whether they were content for these to be attributable or not.

### INTERVIEW CONTENT, CONDUCT AND THEMATIC ANALYSIS

The study was qualitative in design. The interview topic guide was informed by literature searches on both high performance team characteristics and healthcare teamwork, and the experience of several authors working in clinical environments and conducting research on teamwork in surgery.

The aim of the interview guide was to explore the beliefs of successful team trainers about how training can produce a strong culture of mutual support and resilience (see Appendix/Table).

Specific questions were included to address the important challenges of team work in clinical settings, the process of effective team formation and how successful trainers maintain team performance under pressure.

Four team members conducted interviews (LE, ES, PM & MI), which lasted up to 60 minutes over Microsoft Teams. Interviewing continued until no new themes were identified. A thematic analysis of the interview material was then conducted^11^. Interviews were recorded and transcribed using a combination of audio recording with verbatim transcription and handwritten notes. Both transcriptions and notes were imported into NVivo12 for analysis. Each transcript was read and a coding frame produced in discussion between the four interviewers. Coding was both deductive, drawing on the team’s prior experience and knowledge of team building, and inductive drawing out perspectives from responses to open questions.

We used the outputs of the thematic analysis to develop an initial set of principles for a healthcare team building approach, and proposals for its implementation. In order to reduce the risks of inappropriate adaptation or simplistic adoption of the messages which emerged from the analysis, we sought views on our proposals from four Human Factors experts with health care experience, as well as conducting discussions within the research team and with clinical staff. The Human Factors experts were sent a summary of the principles and our proposals and took part in a structured discussion supervised by MS. We used these discussions to identify parts of the proposals which were likely to prove difficult to implement in hospital care and modify them accordingly.

## RESULTS

### INTERVIEWEES

We conducted 14 interviews, 8 with sports coaches/key players and 6 with military leaders and trainers. 4 interviewees were female, or coached female teams. 2 interviewees had healthcare experience. Interviewees came from a range of team sports and from all three branches of the UK armed services. Several had a high public profile and/or an outstanding reputation based on past success (See Table 1). Three declined to be personally identified but supplied verbatim quotes anonymously.

**Table 1.**
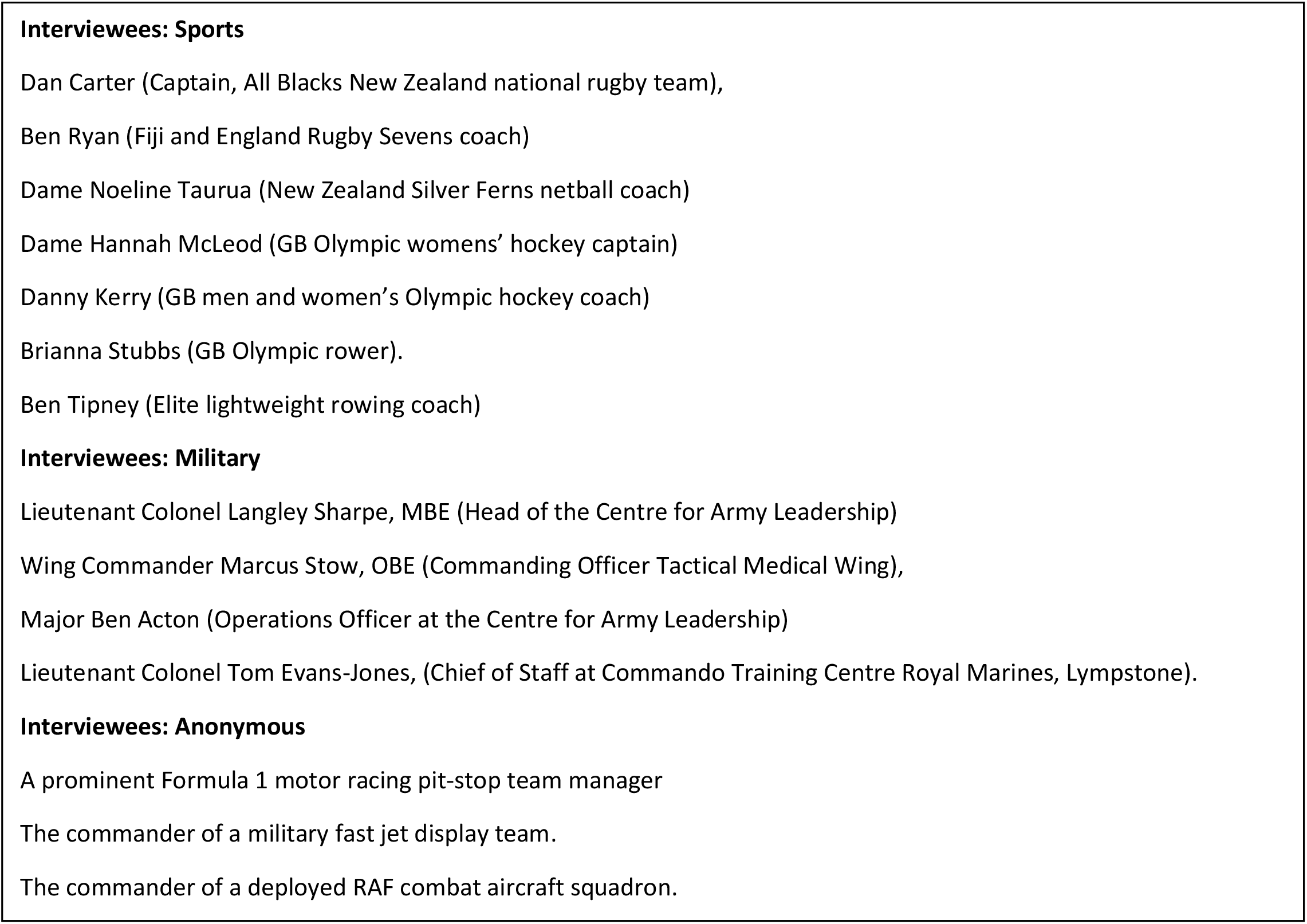
Interviewees.

### THEMES

The six themes we identified are shown in Table 2, with their sub-themes where relevant. There was strong convergence of opinion amongst the interviewees from the military and sporting cohorts about key aspects of team development, with similar responses to the core questions in the topic guide. These included the need for co-design of team vision and purpose by the members, and for creating and maintaining a collective and inclusive culture in which everyone felt safe; The need for frequent intensive training or practice together; clear communication with shared goals; developing an attitude of embracing pressure; informally enforcing team values via experienced members; expecting pro-active anticipation and support for each other, and learning from mistakes instead of condemning individuals.

**Table 2.** Themes and sub-themes.

| Name |
| --- |
| **Communication** |
| **Culture** |
| **Hierarchy** |
| Psychological safety |
| leadership |
| sub-teams |
| **Identity** |
| shared vision |
| socialisation |
| standards and values |
| codes of conduct, mission statements |
| Symbols, rituals, mottos |
| **Building & Maintaining teamwork** |
| continuity stability, team turnover |
| cultural awareness |
| motivation |
| practice and review |
| respect |
| support networks |
| **Embracing pressure** |
| dealing with conflict |
| dealing with failure and mistakes |
| debriefs |
| embracing pressure |

### COMMUNICATION

Encouraging and modelling clear communication with shared goals was emphasised by interviewees, as was the recommendation that activities requiring co-operation and communication should be a core part of regular training. Most teams had codes, acronyms or ways of clearly delivering core information under stress.

#### QUOTES

> *“Establish a safe communication system – learn their names, knowledge and skills - rules are this – speak to me at any time, no-one is to shout, reflect instructions, update. I as a leader will obey the rules, I’m fallible too” MS, RAF*
>
> *“What is important here is working to ensure that every sub-team is well represented and heard*.*” F1 pit stop team director*
>
> *“giving everyone a voice in a no-blame environment” HM*

### CULTURE

Interviewees stressed both the importance of developing and maintaining a positive, inclusive and safe culture in which pro-active mutual support was expected, and the importance of experienced team members in maintaining this culture and passing it on. Several referred to the development of intuition amongst team members who understood instinctively what was needed by others and automatically moved to provide it.

#### QUOTES

> “*There is a team culture but it’s very welcoming to other cultures and as these are embraced it becomes the teams biggest strength – like strings that twist together into a rope*.*” DC, Rugby player/coach*.
>
> “*Senior players play a specific role – they need to behave in a worthy manner that meets the values that are set out. They need right behaviours and to do what they say. Poor leadership will permeate through to junior players*.*” NT, Netball coach*.
>
> *“We adhere to the Pegasus code, which is a shared ethos and vision with a long history to it.” BA*,
>
> *Army School of Leadership*.

### HIERARCHY

None of the coaches questioned the need for hierarchy, but all agreed that leadership needed to be relaxed and confident enough for all team members to feel safe to question authority, air their views and feelings, and make suggestions, including speaking up when they felt something was wrong.

Sub-teams with special functions needed to be firmly embedded in the culture and not excluded or permitted to develop attitudes which excluded others.

#### QUOTES

> *“There is no hierarchy of seniority amongst players – they lead and set culture by example and emphasise the importance of an equal hierarchy when it comes to off field eg: cleaning up after a match. Gratitude, humility, everyone treated the same”. DC, Rugby player/coach*.
>
> *“Hierarchy can stifle initiative and communication”* LS
>
> “*I placed emphasis on ‘guard rails’ – a range for standards where everyone understood the parameters of behaviour. These were established by the players who took ownership of them and were champions of them. For instance a good value system may include values such as respect with the behaviour being ‘turning up on time for practice’. Therefore if players had to be reprimanded on any of these points it wasn’t personal but referred to the value they had all agreed on*.*” BR rugby coach*

### IDENTITY

A range of issues were grouped under this important theme. All interviewees stressed the need for common values and vision. Several stressed the need for members of a newly formed (or reformed) team to define their values together, without external pressure. Interviewees agreed that this required significant time together, and that discussions should be structured so that pre-formed groupings and hierarchies did not compartmentalise or dominate them. Regular team socialisation was important according to most coaches, as a way to strengthen links between team members and encourage mixing between members of different seniority, cultural background and team function. Most coaches believed that symbols and statements of belonging and or team purpose, such as uniforms, emblems, mottoes and mission statements, were useful.

#### QUOTES

> “*Make them believe they belong, an important value. Shared experience. SHARP FOCUS, SHARED EXPERIENCE, CREATING ENVIRONMENT of experiences and hardships. TEJ, Royal Marines*
>
> *“the team is always more important than the individual, so every decision you make you need to put*
>
> *the team first. That’s one of the biggest values”. DC, rugby player/coach*.
>
> “*Symbols of identity are important, for instance a RM green beret is a rite of passage, singles you out, makes you part of the club that has values of excellence, humility, cheerfulness under adversity. Have to live them*.*” TEJ, Royal Marines*.
>
> *“Getting together and knowing and understanding the people on the team is very important*.*”* BA

### BUILDING AND MAINTAINING TEAMWORK

Interviewees were asked about training methods, and how they dealt with specific challenges to team cohesion. All agreed on the importance of frequent intensive training or practice together, with feedback or review of performance. In discussing the difficulties of frequent turnover of team membership, interviewees stressed how important longstanding team members were in maintaining the culture when turnover was high. Encouraging the development of support networks and the use of buddying and mentoring were mentioned by several interviewees. Leadership had an important role in maintaining motivation, mutual respect and cultural awareness. Most coaches would not tolerate players who disregarded the team ethos or appeared to put their own interests ahead of those of the team.

#### QUOTES

> *“We deliberate practice discussing about contextual randomness and the principle of adaptation. We train in different scenarios which increase the ability to classify problems and produce solutions especially in high pressure settings. Staying calm and self-regulating emotions is a major part of the*
>
> *training”*. DK, Field hockey Coach
>
> *“even the best have multiple stages to go through – “age and stage” idea. Older players need to*
>
> *support younger ones with both love and tough love*.*”. NT, netball coach*.
>
> *“Mentorship is key for encouraging ethos of unit in understanding, reflection, debriefing and own failings*.*” MS, RAF*.
>
> *“We have a “no dickheads” rule in the All Blacks” DC rugby player/coach*

### EMBRACING PRESSURE

A key concept amongst successful coaches was encouraging team members to engage with and take strength from pressure rather than fearing or avoiding it. One important aspect of this was to stress learning from failure rather than blaming individuals or groups. Debriefing sessions should be carefully managed to achieve this. Trust between team members was seen as a fundamental part of a successful culture. Celebrating success is important as a way of building confidence that pressure can be overcome. Conflict arises at times within all teams, and it is important to have agreed ways of resolving this without harming the team. Leaders need to know their team well to anticipate and avert problems.

#### QUOTES

> “*challenging them to take them out of that safe (comfort) zone can help resilience*.*” Falklands*

#### Officer RAF

> *“The next 4 years was spent creating a culture of trying to embrace pressure. This included growth mindset and training sessions which recreated high pressure environments with high intensity*.
>
> *Change your mindset, walk towards pressure - It’s a privilege to have pressure in your life, in high performance it’s a sign you’re on the verge of greatness*.*” DC, rugby player/coach*
>
> *“Survivorship bond”* and *“Excellence is something you chase”* TEJ

### PRINCIPLES AND PROSPOSALS FOR DEVELOPMENT OF A TEAM STRENGTHENING PROGRAMME

The principles for a healthcare team strengthening programme based on the results of thematic analysis are outlined in Table 3. These were used to create a set of initial proposals for a healthcare setting, alongside the contextual challenges highlighted by our Human Factors experts and frontline staff advisors in adapting them to the healthcare environment (Table 4).

**Table 3.**
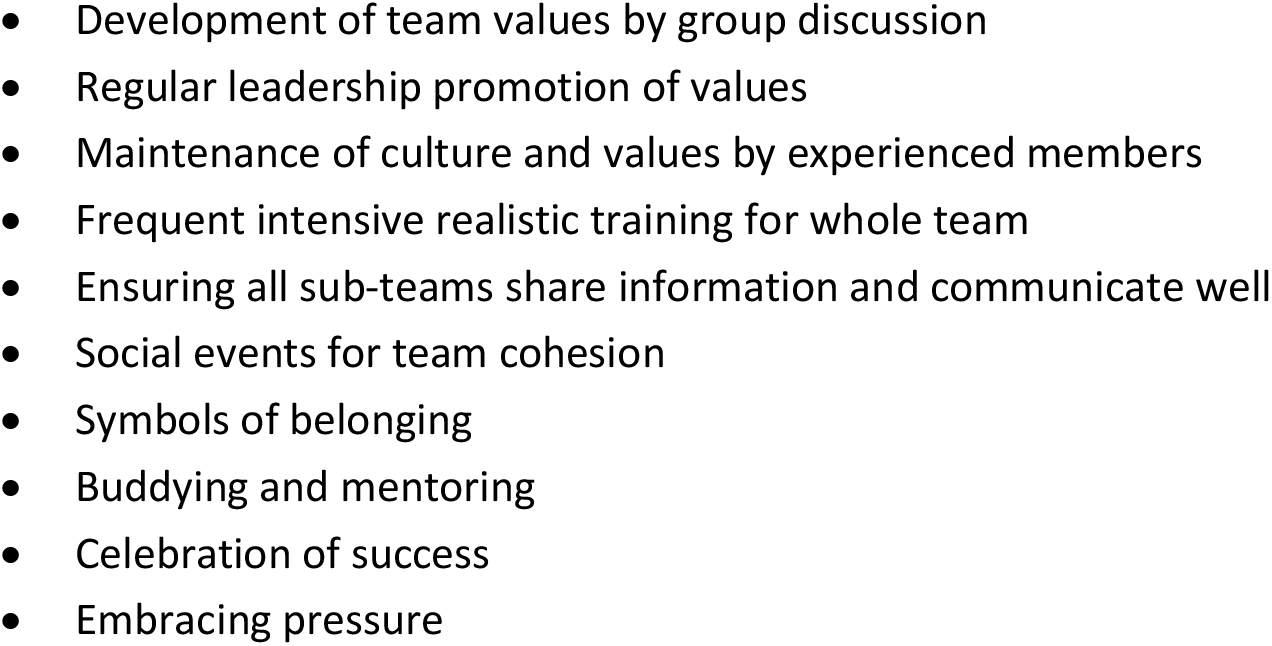
Principles for Team Strengthening from Thematic Analysis.

**Table 4.** Proposal and Consideration of Contextual Challenges.

| Proposals | Challenges |
| --- | --- |
| 1. “Awayday” events to define team values: involving most of the team both nursing and medical | A major logistical operation in terms of rota management. Two identical 3-4 hour events close together, each with about 30% of a Unit’s staff is a possible solution |
| 2. Leaders routinely promote and remind team of their values, including embracing pressure | Not customary in many healthcare settings; leaders may need training: sensitive development required to find the right approach for context |
| 3. Experienced staff to maintain values and instil them in new recruits | This is a natural role for these staff, so likely to be effective – if the experienced staff can first be convinced |
| 4. Frequent realistic intensive low tech in situ crisis simulation training | Logistically very difficult: May need to substitute observed practice with feedback on team interaction and performance instead |
| 5. Regular “huddles” for Dr/nurse communication about seriously ill patients | Leadership required to overcome scheduling difficulties and differences in focus |
| 6. Regular informal team social events | Societal change has made people less willing to socialize with work colleagues after hours: may need to be conducted within work environments |
| 7. Uniforms, emblems, mottoes, badges to emphasise belonging | Need to be generated by staff groups to have credibility: There may be differences in how |
|  | men and women view the value of these symbols |
| 8. Buddying & Mentoring | Already in place for NHS nurses – culturally unfamiliar for Drs, but no obvious barriers |
| 9. Celebrating team success or outstanding performance – appreciative enquiry boards and messaging | Existing structures can be adopted and enhanced, but Unit-level systems are needed for best effect |
| 10. Systems for encouraging mutual support using cards to message within the team | Needs experimental testing: may be uncomfortable for some teams. |

- Development of team values by group discussion
- Regular leadership promotion of values
- Maintenance of culture and values by experienced members
- Frequent intensive realistic training for whole team
- Ensuring all sub-teams share information and communicate well
- Social events for team cohesion
- Symbols of belonging
- Buddying and mentoring
- Celebration of success
- Embracing pressure

The most obvious difference between the elite sports/military contexts and healthcare was the focus on regular, intense and realistic training as a team. In both military and sports contexts this is feasible because of the high ratio of training time to time spent delivering the activity trained for. This ratio is reversed in healthcare, so different approaches will be required, such as brief observations of teams in action with feedback on teamwork performance – and empirical evidence will be needed to show whether these adaptations are successful.

## DISCUSSION

There is a need for a more sophisticated understanding of teamwork both in the surgical arena and in the wider healthcare setting, as part of a re-assessment of our approaches to making healthcare safer and more reliably effective. The idea that good teamwork is helpful in healthcare, as in other workplaces where skilled specialist workers need to cooperate, is not controversial, and it is generally accepted that it is likely to improve outcomes for patients. Whilst there are some differences in the descriptions of good teamwork in relevant literature^12,13^ there is a strong consensus around the nature of the core elements. These include effective co-operation, a lack of psycho-social barriers to communication and a group commitment to pro-active mutual support in achieving specific goals together. How teamwork may be purposively strengthened is however less well established.

The CRM-based model widely used in healthcare concentrates on making staff aware of their cognitive, perceptual and psychosocial limitations and proposing defences against these, but pays less attention to the positive aspects of building the team as a social construct, which are at the core of the work of teamwork experts in high-stakes environments such as elite sports and warfare^14-17^. CRM-inspired models implicitly assume a relatively simple, predictable, highly reliable and well-resourced work system, very different from the context of much secondary healthcare work internationally, and consequently propose algorithmic “Safety 1” constructs such as checklists and standard operating procedures as the basis for improving performance. By focusing principally on staff limitations, this approach transgresses one of the main principles of effective team development, which is to build and maintain a positive attitude around the capacity of the team to succeed^7,18^. The principles espoused by our interviewees seemed to align more closely with resilience concepts underlying the “Safety 2” approach to Human Factors which has been gaining popularity^19^. The most persuasive argument for considering alternatives, however, is the weak empirical evidence of an impact on patient outcomes from current methods.

These observations stimulated our interest in exploring alternative models for improving team cohesiveness, proactive mutual support and resilience in real life healthcare settings. Elite sports and military coaches appeared to us to have a credible claim to expertise in developing highly functional mutually supportive teams which can operate successfully in high pressure situations.

Thematic analysis of our interviews with such coaches confirmed a close affinity between the viewpoints of sports and military coaches around a clear set of potential principles for the development of training courses. The themes identified encompassed a range of issues which are familiar to students of teamwork in healthcare and the explanations for the success of their methods provided by our interviewees were clear, coherent and credible. Some of the most important components of team building, such as internal development of team values through discussion and practice seemed easily transferable to healthcare settings, whilst others, such as frequent brief intensive team training sessions clearly posed a challenge because of the very different context. To minimise the risks of inappropriate “read across” between contexts, we engaged both healthcare Human Factors experts and active clinical staff to review our findings, and critique our proposed training model. The modifications proposed in Table 4 represent a pragmatic “best guess” about how the principles should be modified to fit the environment, and require empirical testing, improvement and validation in live clinical environments. We intend to pilot the modified proposals in our current research programme, which aims to develop Human Factors interventions to improve outcomes after complications from emergency abdominal surgery. Both broadly based^16^ and specifically sports literature oriented^20^ systematic reviews have shown a strong correlation between improvements in process and performance and the adoption of new training schemes aimed at improving cohesion and cooperation, suggesting that the approach merits serious consideration and testing in healthcare.

Our selection of experts was purposive, and we succeeded in interviewing experts with demonstrable track records of success in team sports and in military units, but the sampling was strongly influenced by availability. In particular the selection was weighted towards male teams and individuals with an Anglo-Saxon cultural background. It is possible that a culturally different selection of experts might have highlighted different themes, leading to different recommendations. Our analysis was mixed i.e. partly inductive and partly deductive, because we believed that we understood some of the most important factors to evaluate in healthcare team development because of our pooled experience working in healthcare. This experience certainly gave us some useful insights, but may have blinded us to others because we have unrecognised biases rooted in our own healthcare cultural attitudes. The training proposals we have derived are based on extrapolation using judgement and experience, and will need empirical testing and iterative improvement before we can expect that an effective programme will emerge. If this can be achieved and a stable, effective programme emerges, formal study will be needed using an appropriate evaluation framework to establish apparent effectiveness in heterogenous populations, and relative effectiveness in a randomised trial.

## Data Availability

The data from this study form part of the data archive for the RESPOND NIHR Programme. These data will be lodged in the University of Oxford data repository (ORA) at the conclusion of the study. Prior to this, data requests should be made to the corresponding author.

## Ethics

The research has ethical approval by the Health Research Authority and IRAS approval from Cambridge East REC (IRAS project ID 270881, REC reference 20/EE/0259).

